# The Accuracy of Healthcare Worker versus Self Collected (2-in-1) Oropharyngeal and Bilateral Mid-Turbinate (OPMT) Swabs and Saliva Samples for SARS-CoV-2

**DOI:** 10.1101/2020.09.17.20197004

**Authors:** Seow Yen Tan, Hong Liang Tey, Ernest Tian Hong Lim, Song Tar Toh, Yiong Huak Chan, Pei Ting Tan, Sing Ai Lee, Cheryl Xiaotong Tan, Gerald Choon Huat Koh, Thean Yen Tan, Chuin Siau

**Author notes:** Corresponding Author (SYT). These authors contributed equally to this work (Seow Yen Tan, Hong Liang Tey, Ernest Tian Hong Lim, Song Tar Toh). GCHK, TYT, CS are Joint Senior Authors.

## Abstract

**Background:** Self-sampling for SARS-CoV-2 would significantly raise testing capacity and reduce healthcare worker (HCW) exposure to infectious droplets personal, and protective equipment (PPE) use.

**Methods:** We conducted a diagnostic accuracy study where subjects with a confirmed diagnosis of COVID-19 (n=401) and healthy volunteers (n=100) were asked to self-swab from their oropharynx and mid-turbinate (OPMT), and self-collect saliva. The results of these samples were compared to an OPMT performed by a HCW in the same patient at the same session.

**Results:** In subjects confirmed to have COVID-19, the detection rates of the HCW-swab, self-swab, saliva, and combined self-swab plus saliva samples were 82.8%, 75.1%, 74.3% and 86.5% respectively. All samples obtained from healthy volunteers were tested negative. Compared to HCW-swab, the detection rates of a self-swab sample and saliva sample were inferior by 8.7% (95%CI: 2.4% to 15.0%, p=0.006) and 9.5% (95%CI: 3.1% to 15.8%, p=0.003) respectively. The combined detection rate of self-swab and saliva had a higher detection rate of 2.7% (95%CI: −2.6% to 8.0%, p=0.321). The sensitivity of both the self-collection methods are higher when the Ct value of the HCW swab is less than 30. The negative correctness of both the self-swab and saliva testing was 100% (95% CI 96.4% to 100%).

**Conclusion:** Our study provides evidence that detection rates of self-collected OPMT swab and saliva samples were inferior to a HCW swab, but they could still be useful testing tools in the appropriate clinical settings.

## Introduction

The current “gold standard” for testing for SARS-CoV-2 requires health care workers to collect a nasopharyngeal (NP) sample from a patient. NP sampling is very uncomfortable for the patient and requires deployment of trained personnel and use of personal protective equipment (PPE) which are in limited supply.

A prior study has shown that a combination of oropharyngeal and anterior nares swabs is equivalent in sensitivity to an NP swab in 190 ambulatory symptomatic patients.^1^ Another study on 236 ambulatory, literate, mostly adult subjects the performance of self-collected nasal and throat swabs was at least equivalent to that of health worker collected swabs for the detection of SARS-CoV-2 and other respiratory viruses.^2^

The international community is actively searching for an even less invasive means of sample collection: saliva. In a recent study by Yale University on 29 subjects,^3^ it was suggested that a large volume sample of saliva collected from COVID-19 inpatients can be more sensitive than NP swabs for SARS-CoV-2 detection, and saliva samples had significantly higher COVID-19 viral titres than NP swabs (p=0.001). Furthermore, the same study showed that sensitivity of COVID-19 in saliva was more consistent throughout extended hospitalization compared to NP swabs.

In addition, there are a number of studies done on saliva testing for COVID-19 which have shown promising results, reporting 91.7%, and 100% positivity in saliva samples of COVID-19 patients.^4,5^ Iwasaki et al found an overall concordance rate of 97.4% for COVID-19 detection with a strong concordance between NP swabs and saliva sampling (κ=0.874) among 66 COVID-19 negative and 10 COVID-19 positive subjects.^6^ Furthermore, a study done by To et al. showed that viral RNA could still be detected in saliva samples in a third of their twenty-three patients 20 days or longer after symptoms onset despite the development of COVID-19 antibodies.^7^ A meta-analysis conducted on 26 saliva studies also showed a positive detection rate of 91%, comparable to the detection rate of 98% from nasopharyngeal swabs.^8^ All these studies had small sample sizes (all <30 COVID-positive subjects) and only one study also sampled COVID-negative subjects.

It is still currently unknown whether self-collected combined OPMT sample, or a self-collected saliva sample is equivalent to a swab done by a health care worker (HCW). Self-collection of samples would reduce very significantly on the reliance of trained personnel to collect samples and ramp up testing capacity. It would also reduce greatly the biosafety risk that is posed to HCWs and help with PPE conservation efforts.

## Materials and methods

### Study design and trial oversight

This was a prospective study involving 401 subjects who were previously tested positive for COVID-19 by RT-PCR, and 100 healthy volunteers. This study was approved by the SingHealth Centralised Institutional Review Board. Written informed consent was obtained from the subjects.

### Participants

The first group consisted of patients who were confirmed to have COVID-19, and who were cared for in either a hospital (Changi General Hospital), or a community care facility (Community Care Facility @ EXPO). Inclusion criteria for the study were patients that were admitted within 3 days prior to recruitment to the study sites. The second group comprised of healthy volunteers who were asymptomatic and well on the day of the study, with no recent COVID-19 exposure. Subjects had to be able to understand and comply with the study instructions.

### Test Procedures

Study subjects underwent three sequential test sample collection procedures within one study episode in the following order:

1. Each subject self-collected a sample combining OP and bilateral MT swabs using a single swab stick;
2. A trained healthcare worker then collected a combined OP and bilateral MT swab using another single swab stick;
3. The subject then self-collected a saliva sample.

Study subjects were shown instructional videos for both the OPMT self-swab and saliva collection prior to commencing the test procedures. There are two main methods of saliva collection that has been described in the literature; one is the drooling method, while the other is the spitting method, which results in collection of oropharyngeal sputum. For this study, we used the spitting method for saliva collection. Synthetic fibre swabs were used for collection of the OP and MT samples by both subject and healthcare worker, and immediately placed in universal transport medium (UTM), while saliva samples were collected using the SAFER-Sample(tm) (by Lucence Diagnostics). All samples were double bagged and stored at room temperature in a chiller bag and transported to assigned laboratory on the same day.

Nucleic acid extraction was performed using PerkinElmer Nucleic Acid Extraction Kits (KN0212) on the Pre-Nat II Automated Workstation (PerkinElmer®, United States), Extraction of swab samples followed the indicated protocol for oropharyngeal swabs, while extraction of saliva samples followed a protocol consisting of pre-liquefaction with dithiothreitol (protocol attached in supplementary materials). R*everse transcription polymerase chain reaction* was performed on the Quantstudio™ 5 Real Time PCR system (Thermo Fisher, United Kingdom) using the PerkinElmer® SARS-CoV-2 Real-time RT-PCR Assay. Any detected Ct value was accepted as a true positive result for SARS-CoV-2 virus.

### Outcomes

The primary objective of the study was to evaluate the accuracy of self-collected (2-in-1) OPMT swabs and self-collected saliva samples for SARS-Cov-2 versus that of HCW-collected (2-in-1) MT and OP swabs. The secondary objective was to evaluate the correlation of PCR Cycle Threshold (Ct) values of self-collected saliva samples and swabs with comparator healthcare worker-collected swabs.

### Sample Size

Firstly, we postulated that OPMT self-swabbing was as accurate as HCW-obtained swabs. An error rate of less than 1% was determined to be of clinical relevance so a sample size of at least 400 subjects was calculated. With the computed sample size of 400 subjects, a non-inferiority could be achieved with at most a 7% difference for OPMT self-swabbing compared to the HCW-obtained swabs. If the study included only subjects who were diagnosed with COVID-19, all positive results would be regarded as true positives. Hence, to address that gap in the form of specificity of self-swabs and saliva testing in the diagnosis of COVID-19, a further study on 100 healthy subjects was conducted. The hypothesis was that with 100% accuracy, the error rate for a false negative was 3.6%.

### Statistical Analysis

All analyses were performed using SPSS 25.0 with statistical significance set at p < 0.05. The estimates for the positivity results of the 3 methods were presented as numbers and percentages. The differences with 95% confidence interval (CI) between self-collection methods and HCW-obtained swabs to assess for non-inferiority was calculated. Sensitivity and specificity of the two self-collection methods were compared with HCW-obtained swabs and results were stratified by Ct values. Spearman’s test was used to assess the correlation of the PCR Ct values across the 3 groups.

## Results

A total of 401 COVID-19 positive and 100 COVID-19 negative subjects who were recruited.

All subjects went through the test procedures - 400 participants were able to provide all 3 samples, while one participant was unable to provide a saliva sample despite a prolonged attempt. All participants tolerated the test procedures well and did not experience any adverse events.

Twenty-seven (6.7%) patients were tested negative across all 3 samples. This may be explained by the fact that they are recovering patients and viral shedding may have ceased at point of testing. Forty-two (10.5%) subjects reported ≥ 1 symptom of acute respiratory infection (ARI) (e.g. fever, cough, rhinorrhoea, sore throat, malaise) on the day of study recruitment while 371 (92.5%) subjects reported being within 7 days from onset of COVID-19 illness.

The detection rates of the HCW swab, self-swab, saliva, and combined self-swab plus saliva samples were 82.8%, 75.1%, 74.3% and 86.5% respectively (**Table 1**). Compared to HCW-swabs, the detection rate was lower for self-swab by 8.7% (95% confidence interval, CI=2.4% to 15.0%, p=0.006) and for saliva samples by 9.5% (95%CI=3.1% to 15.8%, p=0.003). When the results of both the self-swab and saliva testing were combined, the detection rate was higher by 2.7% (95%CI=-2.6% to 8.0%, p = 0.321) but this was not statistically significant.

**Table 1.** Detection rates of various modalities in all subjects.

|  | **HCW Swab** | **Self-Swab** | **Saliva** | **Self-Swab + Saliva** |
| --- | --- | --- | --- | --- |
| Count | 336 | 301 | 297 | 347 |
| Percentage | 83.8% | 75.1% | 74.3% | 86.5% |
| 95% CI | 79.8% - 87.3% | 70.1% - 79.2% | 69.7% - 78.5% | 82.8% - 89.7% |

The sensitivities of the self-saliva, saliva and combined self-swab plus saliva testing, when compared to the HCW swab were 83.6%, 80.6% and 92.3% respectively (**Table 2**).

**Table 2.** Sensitivity of Self-Swab, Saliva, and combination of Self-Swab and Saliva in all subjects (n=401) compared to HCW Swab.

|  | **Self-swab** | **Saliva** | **Self-swab + Saliva** |
| --- | --- | --- | --- |
| Sensitivity (95% CI) | 83.6%<br>(79.2 - 87.4) | 80.6%<br>(75.9 - 84.7) | 92.3%<br>(88.9 - 94.9) |

Using the Ct values of HCW swabs as reference, 3 categories of Ct values (i.e. <25, 25-30 and >30) were studied. It was observed that the sensitivity of self-swab (**Table 3**) and saliva testing (**Table 4)** performed better at the lower Ct values, suggesting that the sensitivity of self-collection methods approaches to that of HCW swab, when the viral load was higher

**Table 3:** Sensitivity of Self-swab, stratified by Ct values of HCW swab.

| **HCW Swab Ct** | **Number of subjects** | **Sensitivity** |
| --- | --- | --- |
| <25 | 60 | 100% (94.0 – 100) |
| 25 – 30 | 81 | 96.3% (89.6 – 99.2) |
| >30 | 195 | 73.3% (66.5 – 79.4) |

**Table 4:**
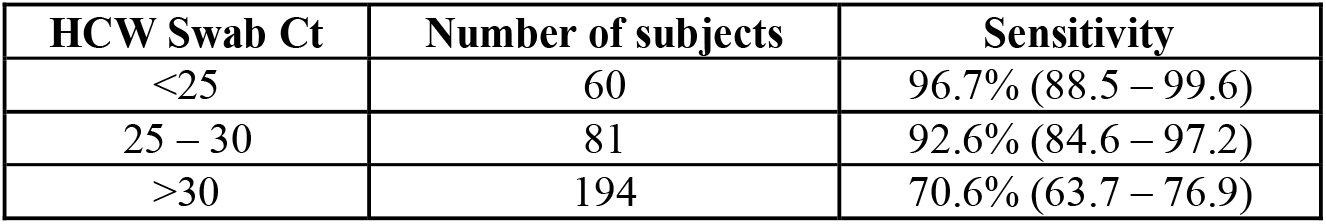
Sensitivity of Saliva, stratified by Ct values of HCW swab.

| **HCW Swab Ct** | **Number of subjects** | **Sensitivity** |
| --- | --- | --- |
| <25 | 60 | 96.7% (88.5 – 99.6) |
| 25 – 30 | 81 | 92.6% (84.6 – 97.2) |
| >30 | 194 | 70.6% (63.7 – 76.9) |

There was a good correlation of PCR Ct values between self-swab and HCW swab (r=0.825, p<0.001) but moderate correlation between saliva samples and HCW swab (r=0.528, p<0.001).

One hundred healthy volunteers were recruited, and all of them were able to provide the 3 required samples. All the samples obtained from the healthy volunteers were tested negative for SARS-CoV2. This implies that the negative correctness was 100% (95% CI 96.4% to 100%) with an error rate of 3.6% for having a false negative.

## Discussion

This study shows that the viral detection rates of a self-sample alone or saliva sample alone is lower than HCW swab. However, the detection rate of a combined self-swab and saliva collection is equivalent to that of a health care worker swab. Another significant finding is that the self-swab and saliva samples have a higher sensitivity with higher viral load which occurs during early illness. The sensitivity of both self-swab and saliva testing drops significantly when the Ct values of the HCW swab is more than 30. A study from Singapore (unpublished data) reported that virus cultures were negative from samples with Ct values >= 30 (i.e. when viral load is low), and the SARS-CoV-2 virus often cannot be isolated or cultured after day 11 of illness.^9^ Our findings corroborate with existing epidemiologic data which indicates that while viral RNA detection may persist in some patients, such persistent RNA detection likely represents non-viable virus and hence, such patients are non-infectious. Thus, the results of this study support the use of self-testing methods as a replacement for HCW swab in the early phase of COVID-19 illness when viral loads are high.

This study included a high proportion of asymptomatic patients who were picked up because of Singapore’s proactive mass screening policy. The combination of self-swab and saliva sampling performed well in these asymptomatic subjects, proving that this strategy has the ability for rapid up-scaling to mass screen patients who are asymptomatic. The study results from the healthy volunteers indicate a low false positive rate with self-collection methods. Thus, self-collection methods may be a useful tool for COVID-19 surveillance in a low prevalence population.

The study team members observed that, despite clear instructions, many subjects still needed guidance with the self-collection methods. For the self-swab, the most commonly encountered problem was subjects needing some guidance in breaking the swab stick. The saliva collection presented a greater challenge to the subjects. The flow of saliva from the funnel into the collection container was not smooth, and the additional step of adding the stabilising fluid required prompting. These necessitated the presence of a trained staff to troubleshoot and ensure that the correct steps are carried out. We believe that these observations are useful in the re-design of collection containers to enhance results and end users’ acceptability. Both the self-swab and saliva collection require dexterity and this would limit its applicability in segments of the population who are not able to do so.

We caution against widespread, unsupervised implementation of self-collection methods. The reliability and effectiveness of self-collection methods may be dependent on social and economic drivers. The test performance may vary, due to factors driving the circumstances. For example, individuals who face a potential loss of income or unemployment if tested positive or travellers having a test done at immigration clearance may deliberately do a suboptimal self-test to influence the test outcome.

Hence, it is important to have designated personnel to supervise the self-collection process. These personnel need not be a HCW and the supervision process will have a lower exposure risk (supervisor can be >1m away from subject), compared to the HCW-swabbing process where a HCW is <1m away and face-to-face with the subject.

The main strength of our study is our large sample size and the inclusion of healthy asymptomatic subjects. A key study limitation however, is that the demographics of the COVID-19-positive population was skewed, consisting solely of male migrant workers, the worst affected group of the pandemic in Singapore, at the time this study was conducted.

## Conclusion

This study demonstrates that while self-collection methods have a detection rate of approximately 75%, it is inferior to the rate obtained by the health care worker administered swab (83.8%). The sensitivity of the self-collection methods is, however, higher and correlates better when Ct values of the HCW swabs are less than 30. The combined results of the saliva and self-swab test achieve a rate detection rate equivalent to that of a health care worker administered swab. The negative correctness of the self-collection methods is 100%. Together with a low false positive rates, we postulate that self-collection methods have their roles in diagnosis in early disease and surveillance screening in low prevalence populations.

## Data Availability

All relevant information are available in the manuscript

## Acknowledgements

We thank all clinical, nursing and allied health staff who provided care for the patients at Changi General Hospital, and Community Care Facility @ EXPO; staff in the Changi General Hospital Clinical Trials & Research Unit for coordinating patient recruitment, logistics management and assistance.

## References

1. LeBlanc JJ, Heinstein C, MacDonald J, Pettipas J, Hatchette TF, Patriquin G. A combined oropharyngeal/nares swab is a suitable alternative to nasopharyngeal swabs for the detection of SARS-CoV-2. J Clin Virol. 2020;128:104442. doi:10.1016/j.jcv.2020.104442

2. Wehrhahn MC, Robson J, Brown S, Bursle E, Byrne S, New D, et al. Self-collection: An appropriate alternative during the SARS-CoV-2 pandemic. J Clin Virol. 2020;128:104417. doi:10.1016/j.jcv.2020.104417

3. Wyllie AL, Fourmier J, Casanovas-Massana A, Campbell M, Tokuyama M, Vijayakumar P, et al. Saliva is more sensitive for SARS-CoV-2 detection in COVID-19 patients than nasopharyngeal swabs. medRxiv 2020 doi: https://doi.org/10.1101/2020.04.16.20067835.

4. To KK, Tsang OT, Yip CC, Chan K, Wu T, Chan JM, et al. Consistent detection of 2019 novel coronavirus in saliva. Clinical Infectious Diseases 2020; 71(15):841–3. doi: https://doi.org/10.1093/cid/ciaa149

5. Azzi L, Carcano G, Gianfagna F, Grossi P, Gasperina DD, Genoni A, et al. Saliva is a reliable tool to detect SARS-CoV-2. J Infect. 2020;81(1):e45–e50. doi:10.1016/j.jinf.2020.04.005.

6. Iwasaki S, Fujisawa S, Nakakubo S, Kamada K, Yamashita Y, Fukumoto T, et al. Comparison of SARS-CoV-2 detection in nasopharyngeal swab and saliva [published online ahead of print, 2020 Jun 4]. J Infect. 2020;81(2):e145–e147. doi:10.1016/j.jinf.2020.05.071.

7. To KK, Tsang OT, Leung WS, Tam AR, Wu T, Lung DC, et al. Temporal profiles of viral load in posterior oropharyngeal saliva samples and serum antibody responses during infection by SARS-CoV-2: an observational cohort study. Lancet Infect Dis. 2020;20(5):565–574. doi:10.1016/S1473-3099(20)30196-1.

8. Czumbel LM, Kiss S, Farkas N, Mandel I, Hegyi I, Nagy A, et al. Saliva as a Candidate for COVID-19 Diagnostic Testing: A Meta-Analysis. medRxiv 2020.05.26.20112565; doi: https://doi.org/10.1101/2020.05.26.20112565.

9. Position Statement from the National Centre for Infectious Diseases and the Chapter of Infectious Disease Physicians, Academy of Medicine, Singapore – 23 May 2020

